## Supplementary information for "Observable Variations in Human Sex Ratio at Birth"

### I Overall SRB Distribution

Figure S1 shows that the distributions of sex ratio at the county level (USA) or the kommun level (Sweden) are very similar, with the US having an overall SRB of $0.5142$ and Sweden $0.5139$.

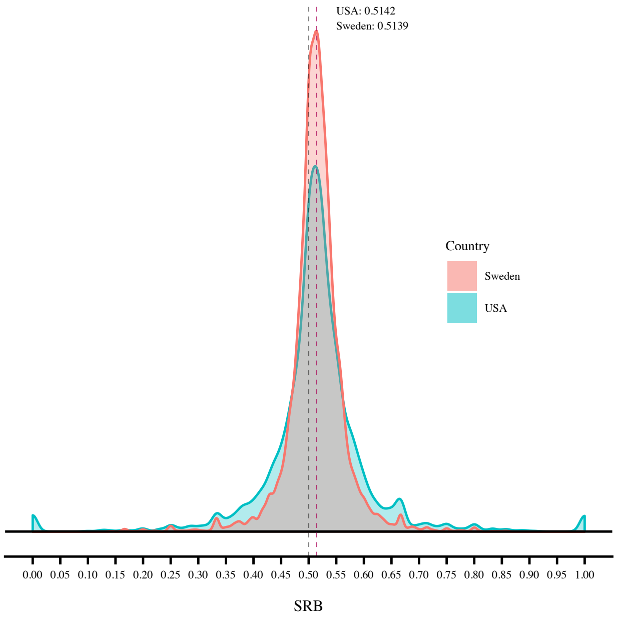


(a) Kernel density estimation (KDE)

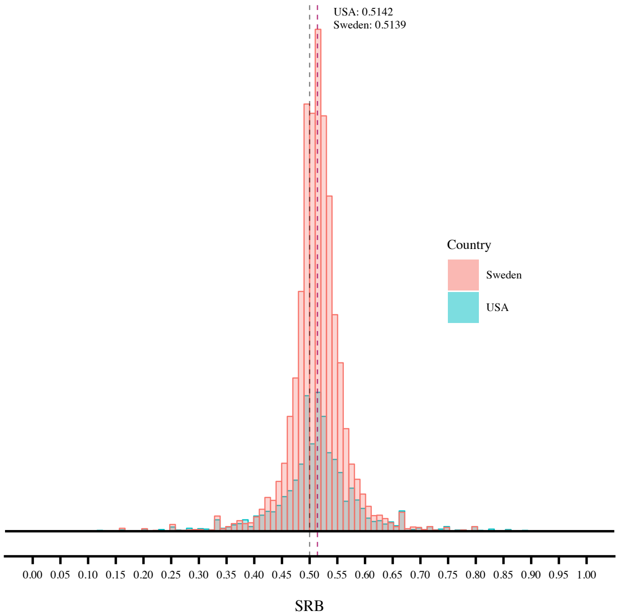


(b) Histogram

Figure S1: Distribution of the SRB in the US and Sweden at the county level (US) or the kommun level (Sweden)

### II Cluster Analysis

Figure S2 shows the dendrogram of the clustering the factors in the US EQI data set by Ward's method (see the Methods section in the main text for more detail). Each red box delimits a statistically significant cluster (at the $95\%$ level), which contains at least $2$ factors.

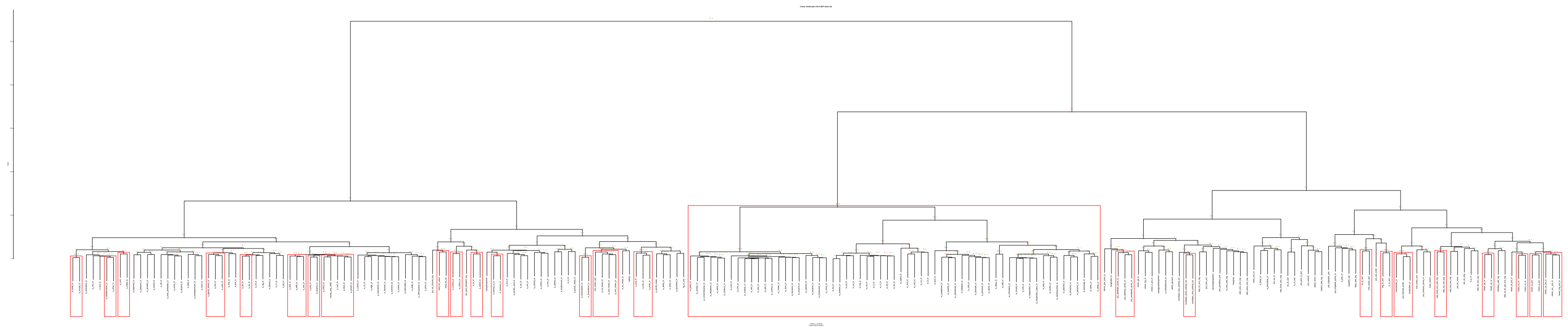

Figure S2: Dendrogram with statistically significant clusters (95% level) in red boxes

### III Regression

Tables S1 and S2, respectively, list all the statistically significant factors (8 for fixed-effect and all 125 for mixed-effect), sorted by decreasing ΔIC, with respect to the null.

| Factor | ΔIC | SE |
|---|---|---|
| cluster_ward_8 | 25.6880 | 7.7901 |
| cluster_ward_1 | 21.5759 | 6.4210 |
| cluster_ward_11 | 20.9930 | 5.1878 |
| cluster_ward_15 | 20.8578 | 4.1703 |
| sewagenpdesperkm | 19.0802 | 4.0902 |
| pct_no_eng | 16.1841 | 4.2216 |
| a_dbp_ln | 13.9700 | 3.8312 |
| pct_mt_10units_log | 10.5599 | 3.3704 |

Table S1: Differences in information criterion (ΔIC) and their standard errors (SE) of individual factors with fixed-effect only. Non-significant factors are omitted.

Table S2: Differences in information criterion (ΔIC) and their standard errors (SE) of individual factors with the random effect at the state level in the US EQI dataset.

| Factor | ΔIC | SE |
|---|---|---|
| pct_rent_occ | 54.2185 | 8.7760 |
| cluster_ward_9 | 52.3907 | 8.8256 |
| mean_pb_ln | 51.7339 | 9.6749 |
| farms_per_acre_ln | 51.6266 | 9.0878 |
| cluster_ward_15 | 51.5511 | 8.5623 |
| pct_mt_10units_log | 51.3259 | 8.5483 |
| rate_food_env_neg | 51.3230 | 9.2907 |
| a_isophorone_ln | 50.9430 | 10.3995 |
| rate_ent_env_log | 50.9259 | 8.8088 |
| a_dbp_ln | 50.4537 | 8.6102 |
| a_mn_ln | 50.2819 | 8.1831 |
| hg_ln_ave | 50.0576 | 8.6584 |
| ryprop | 49.6911 | 9.4066 |
| a_n2h2_ln | 49.5381 | 8.4877 |
| pct_vac_units | 49.3265 | 8.5257 |
| fatal_rate_log | 49.0348 | 8.3392 |
| cluster_ward_25 | 48.6212 | 9.8838 |
| indnpdesperkm | 48.5428 | 10.2509 |
| a_biphenyl_ln | 48.4603 | 8.6128 |
| a_cn_ln | 48.1418 | 9.7888 |
| cluster_ward_10 | 47.7198 | 8.7054 |
| work_out_co | 47.4916 | 8.9555 |
| mean_fe_pct_ln | 47.2117 | 8.0750 |
| cluster_ward_19 | 47.1722 | 8.5983 |
| cluster_ward_26 | 47.1691 | 8.7901 |
| a_quinoline_ln | 47.1477 | 9.6906 |
| a_sb_ln | 47.0255 | 8.3046 |
| cluster_ward_8 | 46.3741 | 8.9328 |

Continued on next page

Table S2: Differences in information criterion ($\Delta$IC) and their standard errors (SE) of individual factors with the random effect at the state level in the US EQI dataset.

| Factor | $\Delta$IC | SE |
|---|---|---|
| med_rooms | 46.3220 | 8.1562 |
| rate_al_pn_gm_env_log | 46.3080 | 8.3765 |
| no3_mean_ave | 46.1843 | 8.8100 |
| a_meoh_ln | 45.9854 | 8.3145 |
| cluster_ward_6 | 45.9191 | 8.6963 |
| d303_percent | 45.8835 | 9.4942 |
| na_ln_ave | 45.0601 | 8.1385 |
| a_co_mean_ln | 44.9827 | 8.5933 |
| a_c6h5cl_ln | 44.6342 | 8.7028 |
| a_me2_phthalate_ln | 44.4614 | 8.0751 |
| a_acrylic_acid_ln | 44.2292 | 9.8383 |
| a_mecl_ln | 44.0207 | 8.4892 |
| cat | 43.9278 | 8.3540 |
| mean_ti_pct_ln | 43.8613 | 8.8678 |
| cluster_ward_18 | 43.8523 | 9.3605 |
| a_c2hcl3_ln | 43.7136 | 8.7911 |
| a_pahpom_ln | 43.6743 | 9.1561 |
| a_benzidine_ln | 43.6200 | 8.4935 |
| rate_civic_env_log | 43.6027 | 8.5611 |
| fungicides_ln | 43.5278 | 8.7614 |
| cluster_ward_3 | 43.5188 | 8.9542 |
| pct_irrigated_acres_ln | 43.4427 | 9.8728 |
| a_p_ln | 43.2101 | 8.2484 |
| to_unit_rate_log | 42.9314 | 8.5853 |
| cluster_ward_2 | 42.8250 | 9.4466 |
| pct_au_ln | 42.8249 | 9.4111 |
| cluster_ward_11 | 42.8243 | 9.0778 |
| a_c3h3n_ln | 42.8093 | 8.4624 |
| a_diesel_ln | 42.7350 | 8.1946 |
| a_etacrylate_ln | 42.7323 | 8.7796 |
| avgofd3_ave | 42.5189 | 8.8904 |
| pct_pub_transport_log | 42.1074 | 8.4142 |
| a_pb_ln | 41.9983 | 8.0841 |
| a_ph3_ln | 41.9038 | 9.0259 |
| a_eox_ln | 41.8434 | 8.9989 |
| pct_unemp | 41.8191 | 7.7675 |
| a_hexane_ln | 41.8044 | 8.0627 |
| pct_no_eng | 41.7541 | 8.2477 |
| a_acrolein_ln | 41.6847 | 8.2058 |
| a_ech_ln | 41.6194 | 8.4322 |
| a_cumene_ln | 41.6100 | 8.2629 |
| a_11dce_ln | 41.5146 | 8.4598 |
| radon_zone | 41.4107 | 8.6552 |
| a_2np_ln | 41.2805 | 9.3701 |
| a_cr_ln | 41.1006 | 8.3650 |
| cluster_ward_13 | 41.0139 | 8.1444 |
| a_pm10_mean_ln | 40.8277 | 8.0920 |
| a_cresol_ln | 40.7598 | 8.2679 |
| rate_trans_env_log | 40.7569 | 8.4773 |
| a_proo_ln | 40.7404 | 8.1918 |

Continued on next page

Table S2: Differences in information criterion (ΔIC) and their standard errors (SE) of individual factors with the random effect at the state level in the US EQI dataset.

| Factor | ΔIC | SE |
|---|---|---|
| per_totpopss_ave | 40.6453 | 8.2117 |
| a_cl_ln | 40.6386 | 8.2518 |
| stormnpdesperkm | 40.5914 | 8.4561 |
| facilities_rate_log | 40.5648 | 9.2041 |
| pct_manure_acres_ln | 40.4699 | 8.8810 |
| cluster_ward_12 | 40.4274 | 9.1367 |
| a_mibk_ln | 40.3786 | 8.7446 |
| a_dehp_ln | 40.3305 | 8.1627 |
| numdays_rain_activity_tot | 40.1392 | 8.3155 |
| cluster_ward_20 | 39.8644 | 8.6439 |
| cl_ln_ave | 39.5243 | 9.2021 |
| a_egly_ln | 39.4799 | 8.2135 |
| mean_hg_ln | 39.4692 | 8.1523 |
| rate_ed_env_log | 39.3155 | 8.7520 |
| a_pm25_mean | 39.1712 | 8.5654 |
| cluster_ward_22 | 39.1668 | 8.3645 |
| a_hg_ln | 39.0513 | 7.8800 |
| cluster_ward_5 | 39.0485 | 8.1282 |
| a_chloroform_ln | 38.9772 | 8.1250 |
| a_cs2_ln | 38.4509 | 7.6905 |
| a_tdi_ln | 38.4377 | 8.9270 |
| per_pswithsw_ave | 38.3795 | 8.1011 |
| a_acetophenone_ln | 37.9548 | 8.6551 |
| violent_rate_log | 37.8928 | 8.8251 |
| a_cd_ln | 37.8764 | 7.9860 |
| a_dbcp_ln | 37.7606 | 8.3128 |
| a_etcl_ln | 37.6943 | 8.3588 |
| a_chloroprene_ln | 37.6642 | 8.4969 |
| cluster_ward_14 | 37.4994 | 8.6078 |
| hwyprop | 37.3042 | 8.4589 |
| a_stryene_ln | 37.2645 | 8.3137 |
| cluster_ward_4 | 37.0974 | 8.2277 |
| cluster_ward_1 | 36.9302 | 8.3875 |
| cluster_ward_16 | 36.9113 | 8.0581 |
| cluster_ward_7 | 36.6871 | 7.8672 |
| rate_hc_env_log | 36.6085 | 8.1125 |
| cluster_ward_23 | 36.5280 | 8.1485 |
| sewagenpdesperkm | 36.0821 | 7.4545 |
| cluster_ward_21 | 35.9375 | 8.7625 |
| a_otoluidine_ln | 35.8421 | 7.9246 |
| a_mma_ln | 35.3633 | 8.1445 |
| a_mehydrazine_ln | 35.1223 | 7.4720 |
| cluster_ward_24 | 35.0831 | 8.0009 |
| pct_pers_lt_pov | 34.9108 | 7.1347 |
| pct_hs_more | 34.8201 | 8.0671 |
| cluster_ward_17 | 34.5813 | 7.9217 |
| a_hcl_ln | 34.1790 | 8.0501 |

### IV Time Series Forecasts

#### IV.1 Monthly

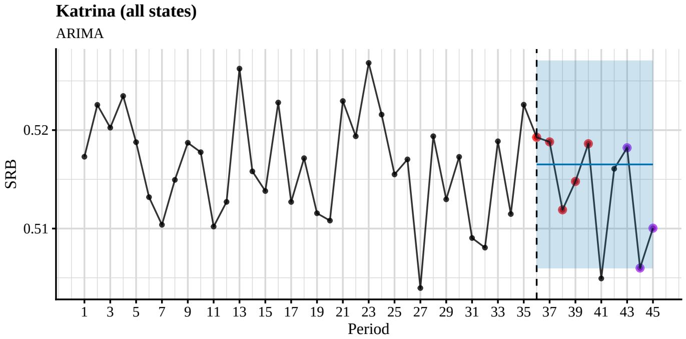


(a) Hurricane Katrina, all states

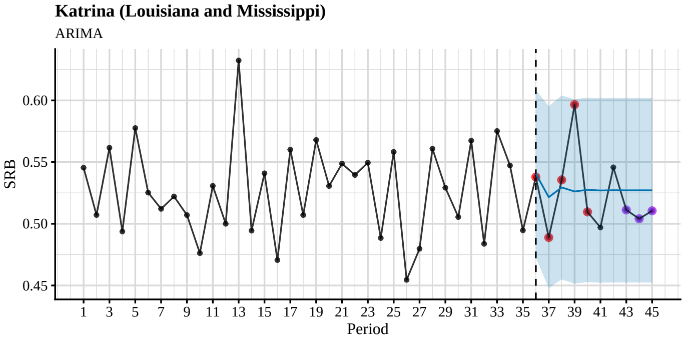


(b) Hurricane Katrina, Louisiana and Mississippi only

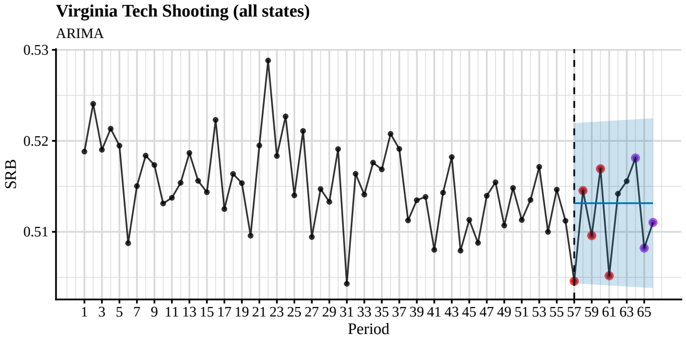


(c) Virginia Tech shooting, all states

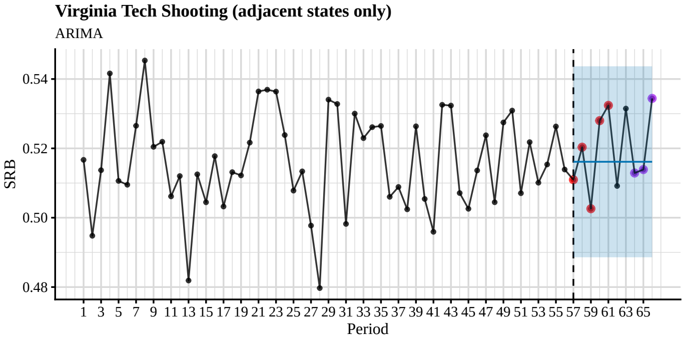


(d) Virginia Tech shooting, adjacent states only

Figure S3: Time series plots and out-of-sample forecasts for SRB data grouped into 28-day periods and fitted with seasonal ARIMA models. The blue shade is the 95% confidence level. The observed SRBs for the first 5 months after the intervention are presented by red dots, whereas the observed SRBs for 7–9 months after the intervention are presented by purple dots. See also Table S3.

| Period | SRB | Lower 95% | Upper 95% |
|---|---|---|---|
| 36 | 0.5193 | 0.5060 | 0.5271 |
| 37 | 0.5188 | 0.5060 | 0.5271 |
| 38 | 0.5119 | 0.5060 | 0.5271 |
| 39 | 0.5148 | 0.5060 | 0.5271 |
| 40 | 0.5186 | 0.5060 | 0.5271 |
| 41(*) | 0.5049 | 0.5060 | 0.5271 |
| 42 | 0.5161 | 0.5060 | 0.5271 |
| 43 | 0.5182 | 0.5060 | 0.5271 |
| 44 | 0.5060 | 0.5060 | 0.5271 |
| 45 | 0.5100 | 0.5060 | 0.5271 |

(a) Hurricane Katrina in all states

| Period | SRB | Lower 95% | Upper 95% |
|---|---|---|---|
| 36 | 0.5379 | 0.4726 | 0.6086 |
| 37 | 0.4889 | 0.4479 | 0.5952 |
| 38 | 0.5356 | 0.4549 | 0.6040 |
| 39 | 0.5966 | 0.4515 | 0.6009 |
| 40 | 0.5096 | 0.4528 | 0.6023 |
| 41 | 0.4970 | 0.4522 | 0.6017 |
| 42 | 0.5458 | 0.4525 | 0.6019 |
| 43 | 0.5112 | 0.4524 | 0.6018 |
| 44 | 0.5041 | 0.4524 | 0.6019 |
| 45 | 0.5105 | 0.4524 | 0.6019 |

(b) Hurricane Katrina in Louisiana and Mississippi

| Period | SRB | Lower 95% | Upper 95% |
|---|---|---|---|
| 57 | 0.5046 | 0.5043 | 0.5220 |
| 58 | 0.5145 | 0.5043 | 0.5220 |
| 59 | 0.5096 | 0.5042 | 0.5221 |
| 60 | 0.5169 | 0.5042 | 0.5221 |
| 61 | 0.5052 | 0.5041 | 0.5222 |
| 62 | 0.5142 | 0.5040 | 0.5222 |
| 63 | 0.5156 | 0.5040 | 0.5223 |
| 64 | 0.5181 | 0.5039 | 0.5224 |
| 65 | 0.5082 | 0.5039 | 0.5224 |
| 66 | 0.5110 | 0.5038 | 0.5225 |

(c) Virginia Tech Shooting in all states

| Period | SRB | Lower 95% | Upper 95% |
|---|---|---|---|
| 57 | 0.5110 | 0.4886 | 0.5437 |
| 58 | 0.5203 | 0.4886 | 0.5437 |
| 59 | 0.5026 | 0.4886 | 0.5437 |
| 60 | 0.5280 | 0.4886 | 0.5437 |
| 61 | 0.5324 | 0.4886 | 0.5437 |
| 62 | 0.5092 | 0.4886 | 0.5437 |
| 63 | 0.5315 | 0.4886 | 0.5437 |
| 64 | 0.5129 | 0.4886 | 0.5437 |
| 65 | 0.5139 | 0.4886 | 0.5437 |
| 66 | 0.5344 | 0.4886 | 0.5437 |

(d) Virginia Tech Shooting in adjacent states

Table S3: Out-of-sample forecasts for the first $10$ months after the intervention using SRB data grouped into $28$-day periods and fitted with seasonal ARIMA models. Any period of which the observed SRB is outside of the $95\%$ confidence level is marked by an asterisk (*).

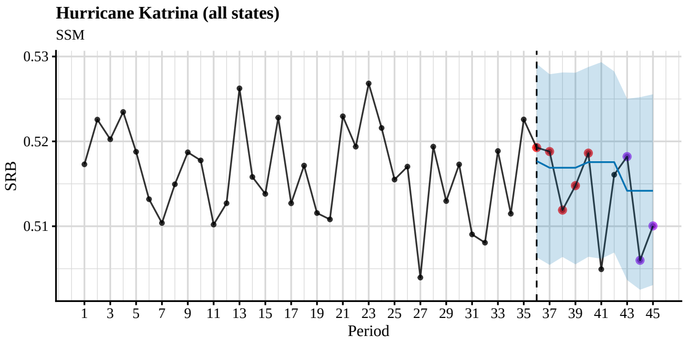


(a) Hurricane Katrina, all states

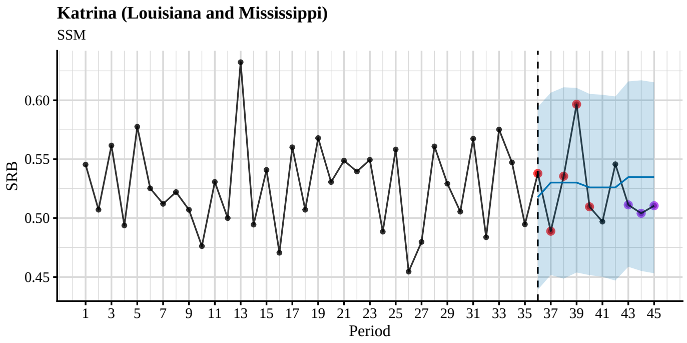


(b) Hurricane Katrina, Louisiana and Mississippi only

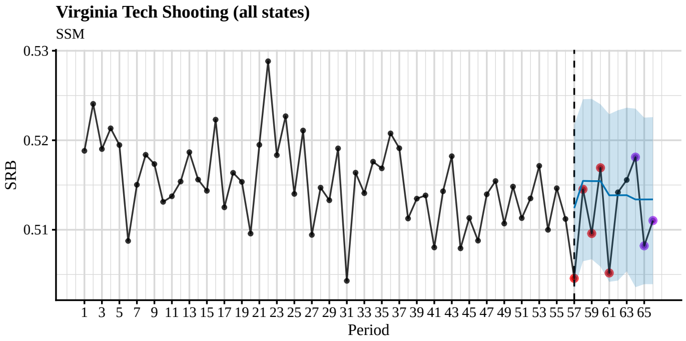


(c) Virginia Tech shooting, all states

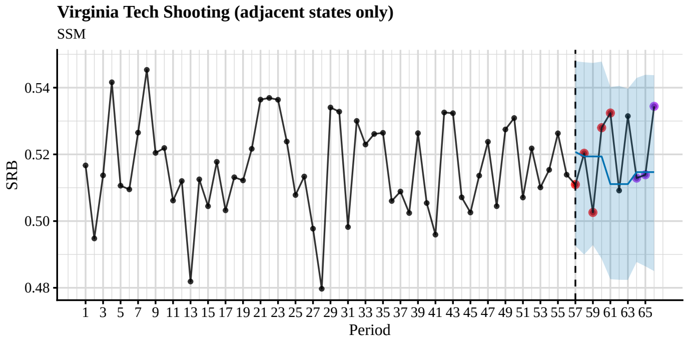


(d) Virginia Tech shooting, adjacent states only

Figure S4: Time series plots and out-of-sample forecasts for SRB data grouped into 28-day periods and fitted with state-space models. The blue shade is the $95\%$ confidence level. The observed SRBs for the first $5$ months after the intervention are presented by red dots, whereas the observed SRBs for 7–$9$ months after the intervention are presented by purple dots. See also Table S4.

| Period | SRB | Lower 95% | Upper 95% |
|---|---|---|---|
| 36 | 0.5193 | 0.5063 | 0.5291 |
| 37 | 0.5188 | 0.5054 | 0.5279 |
| 38 | 0.5119 | 0.5064 | 0.5281 |
| 39 | 0.5148 | 0.5055 | 0.5281 |
| 40 | 0.5186 | 0.5064 | 0.5288 |
| 41 | 0.5049 | 0.5062 | 0.5293 |
| 42 | 0.5161 | 0.5069 | 0.5282 |
| 43 | 0.5182 | 0.5037 | 0.5250 |
| 44 | 0.5060 | 0.5025 | 0.5252 |
| 45 | 0.5100 | 0.5031 | 0.5255 |

(a) Hurricane Katrina in all states

| Period | SRB | Lower 95% | Upper 95% |
|---|---|---|---|
| 36 | 0.5379 | 0.4392 | 0.5944 |
| 37 | 0.4889 | 0.4517 | 0.6065 |
| 38 | 0.5356 | 0.4486 | 0.6111 |
| 39 | 0.5966 | 0.4538 | 0.6105 |
| 40 | 0.5096 | 0.4516 | 0.6054 |
| 41 | 0.4970 | 0.4502 | 0.6047 |
| 42 | 0.5458 | 0.4470 | 0.6033 |
| 43 | 0.5112 | 0.4587 | 0.6160 |
| 44 | 0.5041 | 0.4551 | 0.6170 |
| 45 | 0.5105 | 0.4532 | 0.6153 |

(b) Hurricane Katrina in Louisiana and Mississippi

| Period | SRB | Lower 95% | Upper 95% |
|---|---|---|---|
| 57 | 0.5046 | 0.5034 | 0.5217 |
| 58 | 0.5145 | 0.5065 | 0.5246 |
| 59 | 0.5096 | 0.5067 | 0.5246 |
| 60 | 0.5169 | 0.5059 | 0.5240 |
| 61 | 0.5052 | 0.5042 | 0.5229 |
| 62 | 0.5142 | 0.5043 | 0.5234 |
| 63 | 0.5156 | 0.5054 | 0.5236 |
| 64 | 0.5181 | 0.5036 | 0.5235 |
| 65 | 0.5082 | 0.5039 | 0.5225 |
| 66 | 0.5110 | 0.5039 | 0.5226 |

(c) Virginia Tech Shooting in all states

| Period | SRB | Lower 95% | Upper 95% |
|---|---|---|---|
| 57 | 0.5110 | 0.4924 | 0.5487 |
| 58 | 0.5203 | 0.4910 | 0.5478 |
| 59 | 0.5026 | 0.4926 | 0.5469 |
| 60 | 0.5280 | 0.4896 | 0.5481 |
| 61 | 0.5324 | 0.4832 | 0.5382 |
| 62 | 0.5092 | 0.4815 | 0.5390 |
| 63 | 0.5315 | 0.4829 | 0.5399 |
| 64 | 0.5129 | 0.4894 | 0.5432 |
| 65 | 0.5139 | 0.4866 | 0.5450 |
| 66 | 0.5344 | 0.4846 | 0.5433 |

(d) Virginia Tech Shooting in adjacent states

Table S4: Out-of-sample forecasts for the first 10 months after the intervention using SRB data grouped into 28-day periods and fitted with state-space models. Any period of which the observed SRB is outside of the 95% confidence level is marked by an asterisk (*).

#### IV.2 Weekly

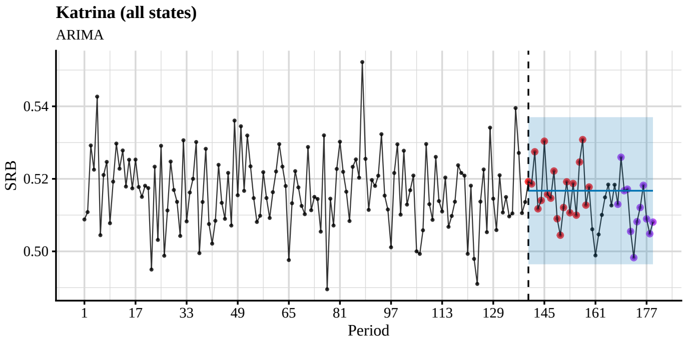


(a) Hurricane Katrina, all states

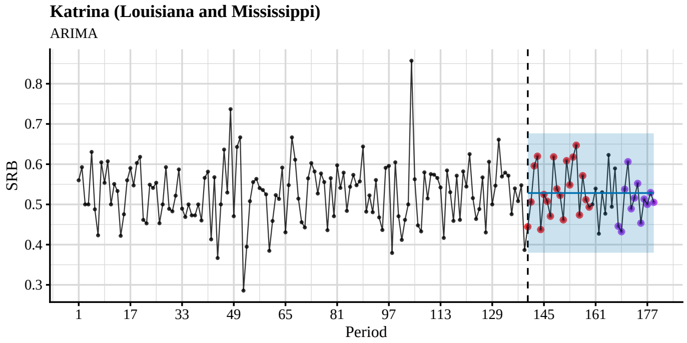


(b) Hurricane Katrina, Louisiana and Mississippi only

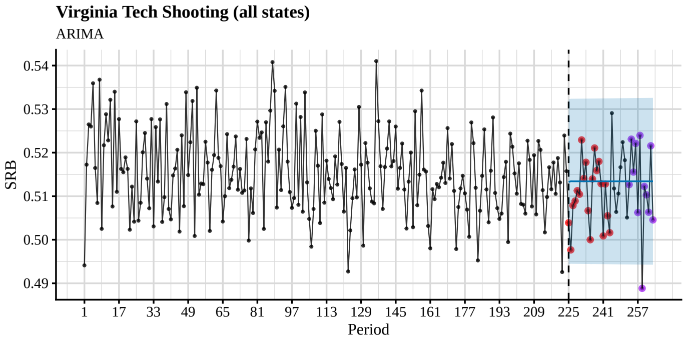


(c) Virginia Tech shooting, all states

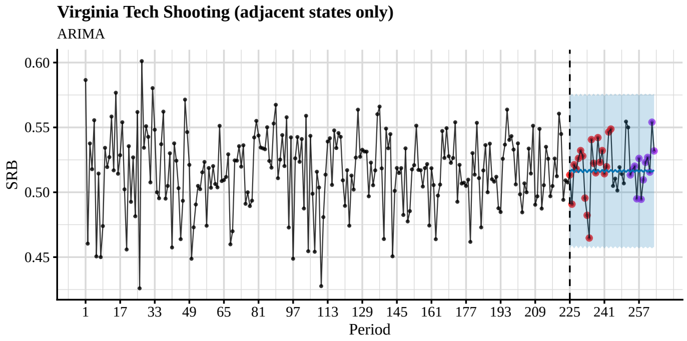


(d) Virginia Tech shooting, adjacent states only

Figure S5: Time series plots and out-of-sample forecasts for SRB data grouped into 28-day periods and fitted with seasonal ARIMA models. The blue shade is the 95% confidence level. The observed SRBs for the first 5 months after the intervention are presented by red dots, whereas the observed SRBs for 7–9 months after the intervention are presented by purple dots. See also Table S5.

| Period | SRB | Lower 95% | Upper 95% |
|---|---|---|---|
| 140 | 0.5192 | 0.4964 | 0.5370 |
| 141 | 0.5186 | 0.4964 | 0.5370 |
| 142 | 0.5275 | 0.4964 | 0.5370 |
| 143 | 0.5117 | 0.4964 | 0.5370 |
| 144 | 0.5140 | 0.4964 | 0.5370 |
| 145 | 0.5304 | 0.4964 | 0.5370 |
| 146 | 0.5157 | 0.4964 | 0.5370 |
| 147 | 0.5147 | 0.4964 | 0.5370 |
| 148 | 0.5222 | 0.4964 | 0.5370 |
| 149 | 0.5090 | 0.4964 | 0.5370 |
| 150 | 0.5045 | 0.4964 | 0.5370 |
| 151 | 0.5121 | 0.4964 | 0.5370 |
| 152 | 0.5192 | 0.4964 | 0.5370 |
| 153 | 0.5106 | 0.4964 | 0.5370 |
| 154 | 0.5187 | 0.4964 | 0.5370 |
| 155 | 0.5100 | 0.4964 | 0.5370 |
| 156 | 0.5246 | 0.4964 | 0.5370 |
| 157 | 0.5308 | 0.4964 | 0.5370 |
| 158 | 0.5128 | 0.4964 | 0.5370 |
| 159 | 0.5177 | 0.4964 | 0.5370 |
| 160 | 0.5061 | 0.4964 | 0.5370 |
| 161 | 0.4989 | 0.4964 | 0.5370 |
| 162 | 0.5047 | 0.4964 | 0.5370 |
| 163 | 0.5101 | 0.4964 | 0.5370 |
| 164 | 0.5149 | 0.4964 | 0.5370 |
| 165 | 0.5184 | 0.4964 | 0.5370 |
| 166 | 0.5127 | 0.4964 | 0.5370 |
| 167 | 0.5184 | 0.4964 | 0.5370 |
| 168 | 0.5130 | 0.4964 | 0.5370 |
| 169 | 0.5260 | 0.4964 | 0.5370 |
| 170 | 0.5167 | 0.4964 | 0.5370 |
| 171 | 0.5171 | 0.4964 | 0.5370 |
| 172 | 0.5055 | 0.4964 | 0.5370 |
| 173 | 0.4983 | 0.4964 | 0.5370 |
| 174 | 0.5082 | 0.4964 | 0.5370 |
| 175 | 0.5121 | 0.4964 | 0.5370 |
| 176 | 0.5182 | 0.4964 | 0.5370 |
| 177 | 0.5089 | 0.4964 | 0.5370 |
| 178 | 0.5049 | 0.4964 | 0.5370 |
| 179 | 0.5081 | 0.4964 | 0.5370 |

(a) Hurricane Katrina in all states

| Period | SRB | Lower 95% | Upper 95% |
|---|---|---|---|
| 140 | 0.4444 | 0.3799 | 0.6767 |
| 141 | 0.5060 | 0.3799 | 0.6767 |
| 142 | 0.5957 | 0.3799 | 0.6767 |
| 143 | 0.6197 | 0.3799 | 0.6767 |
| 144 | 0.4375 | 0.3799 | 0.6767 |
| 145 | 0.5246 | 0.3799 | 0.6767 |
| 146 | 0.5077 | 0.3799 | 0.6767 |
| 147 | 0.4706 | 0.3799 | 0.6767 |
| 148 | 0.6182 | 0.3799 | 0.6767 |
| 149 | 0.5385 | 0.3799 | 0.6767 |
| 150 | 0.5224 | 0.3799 | 0.6767 |
| 151 | 0.4615 | 0.3799 | 0.6767 |
| 152 | 0.6087 | 0.3799 | 0.6767 |
| 153 | 0.5484 | 0.3799 | 0.6767 |
| 154 | 0.6176 | 0.3799 | 0.6767 |
| 155 | 0.6471 | 0.3799 | 0.6767 |
| 156 | 0.4737 | 0.3799 | 0.6767 |
| 157 | 0.5714 | 0.3799 | 0.6767 |
| 158 | 0.5116 | 0.3799 | 0.6767 |
| 159 | 0.4933 | 0.3799 | 0.6767 |
| 160 | 0.5000 | 0.3799 | 0.6767 |
| 161 | 0.5393 | 0.3799 | 0.6767 |
| 162 | 0.4271 | 0.3799 | 0.6767 |
| 163 | 0.5301 | 0.3799 | 0.6767 |
| 164 | 0.4769 | 0.3799 | 0.6767 |
| 165 | 0.6230 | 0.3799 | 0.6767 |
| 166 | 0.4941 | 0.3799 | 0.6767 |
| 167 | 0.5895 | 0.3799 | 0.6767 |
| 168 | 0.4458 | 0.3799 | 0.6767 |
| 169 | 0.4321 | 0.3799 | 0.6767 |
| 170 | 0.5376 | 0.3799 | 0.6767 |
| 171 | 0.6061 | 0.3799 | 0.6767 |
| 172 | 0.4891 | 0.3799 | 0.6767 |
| 173 | 0.5158 | 0.3799 | 0.6767 |
| 174 | 0.5521 | 0.3799 | 0.6767 |
| 175 | 0.4535 | 0.3799 | 0.6767 |
| 176 | 0.5132 | 0.3799 | 0.6767 |
| 177 | 0.5000 | 0.3799 | 0.6767 |
| 178 | 0.5294 | 0.3799 | 0.6767 |
| 179 | 0.5052 | 0.3799 | 0.6767 |

(b) Hurricane Katrina in Louisiana and Mississippi

| Period | SRB | Lower 95% | Upper 95% |
|---|---|---|---|
| 225 | 0.5039 | 0.4943 | 0.5322 |
| 226 | 0.4977 | 0.4945 | 0.5324 |
| 227 | 0.5079 | 0.4945 | 0.5324 |
| 228 | 0.5089 | 0.4945 | 0.5324 |
| 229 | 0.5113 | 0.4945 | 0.5324 |
| 230 | 0.5104 | 0.4945 | 0.5324 |
| 231 | 0.5229 | 0.4944 | 0.5324 |
| 232 | 0.5141 | 0.4944 | 0.5324 |
| 233 | 0.5178 | 0.4944 | 0.5324 |
| 234 | 0.5067 | 0.4944 | 0.5324 |
| 235 | 0.5000 | 0.4944 | 0.5324 |
| 236 | 0.5140 | 0.4944 | 0.5324 |
| 237 | 0.5211 | 0.4944 | 0.5324 |
| 238 | 0.5159 | 0.4944 | 0.5324 |
| 239 | 0.5180 | 0.4944 | 0.5324 |
| 240 | 0.5129 | 0.4944 | 0.5325 |
| 241 | 0.5009 | 0.4944 | 0.5325 |
| 242 | 0.5128 | 0.4944 | 0.5325 |
| 243 | 0.5055 | 0.4944 | 0.5325 |
| 244 | 0.5016 | 0.4944 | 0.5325 |
| 245 | 0.5291 | 0.4944 | 0.5325 |
| 246 | 0.5118 | 0.4944 | 0.5325 |
| 247 | 0.5064 | 0.4944 | 0.5325 |
| 248 | 0.5106 | 0.4944 | 0.5325 |
| 249 | 0.5167 | 0.4943 | 0.5325 |
| 250 | 0.5224 | 0.4943 | 0.5325 |
| 251 | 0.5183 | 0.4943 | 0.5325 |
| 252 | 0.5051 | 0.4943 | 0.5325 |
| 253 | 0.5127 | 0.4943 | 0.5325 |
| 254 | 0.5231 | 0.4943 | 0.5325 |
| 255 | 0.5155 | 0.4943 | 0.5325 |
| 256 | 0.5221 | 0.4943 | 0.5325 |
| 257 | 0.5063 | 0.4943 | 0.5325 |
| 258 | 0.5240 | 0.4943 | 0.5325 |
| 259(*) | 0.4888 | 0.4943 | 0.5326 |
| 260 | 0.5122 | 0.4943 | 0.5326 |
| 261 | 0.5103 | 0.4943 | 0.5326 |
| 262 | 0.5063 | 0.4943 | 0.5326 |
| 263 | 0.5216 | 0.4943 | 0.5326 |
| 264 | 0.5046 | 0.4943 | 0.5326 |

(c) Virginia Tech Shooting in all states

| Period | SRB | Lower 95% | Upper 95% |
|---|---|---|---|
| 225 | 0.5134 | 0.4567 | 0.5739 |
| 226 | 0.4908 | 0.4589 | 0.5761 |
| 227 | 0.5212 | 0.4567 | 0.5739 |
| 228 | 0.5179 | 0.4589 | 0.5761 |
| 229 | 0.5261 | 0.4567 | 0.5740 |
| 230 | 0.5322 | 0.4588 | 0.5761 |
| 231 | 0.5280 | 0.4567 | 0.5741 |
| 232 | 0.4955 | 0.4587 | 0.5761 |
| 233 | 0.4822 | 0.4567 | 0.5741 |
| 234 | 0.4647 | 0.4587 | 0.5761 |
| 235 | 0.5406 | 0.4567 | 0.5742 |
| 236 | 0.5223 | 0.4586 | 0.5761 |
| 237 | 0.5150 | 0.4567 | 0.5743 |
| 238 | 0.5421 | 0.4585 | 0.5761 |
| 239 | 0.5231 | 0.4567 | 0.5743 |
| 240 | 0.5322 | 0.4585 | 0.5761 |
| 241 | 0.5143 | 0.4567 | 0.5744 |
| 242 | 0.5197 | 0.4584 | 0.5761 |
| 243 | 0.5465 | 0.4567 | 0.5744 |
| 244 | 0.5487 | 0.4584 | 0.5761 |
| 245 | 0.5049 | 0.4567 | 0.5745 |
| 246 | 0.5105 | 0.4583 | 0.5761 |
| 247 | 0.5014 | 0.4567 | 0.5745 |
| 248 | 0.5193 | 0.4583 | 0.5761 |
| 249 | 0.5141 | 0.4567 | 0.5746 |
| 250 | 0.5068 | 0.4583 | 0.5761 |
| 251 | 0.5545 | 0.4567 | 0.5746 |
| 252 | 0.5500 | 0.4582 | 0.5761 |
| 253 | 0.5133 | 0.4567 | 0.5746 |
| 254 | 0.5172 | 0.4582 | 0.5761 |
| 255 | 0.5201 | 0.4567 | 0.5747 |
| 256 | 0.4949 | 0.4581 | 0.5761 |
| 257 | 0.5261 | 0.4567 | 0.5747 |
| 258 | 0.4946 | 0.4581 | 0.5761 |
| 259 | 0.5096 | 0.4567 | 0.5748 |
| 260 | 0.5231 | 0.4581 | 0.5761 |
| 261 | 0.5269 | 0.4568 | 0.5748 |
| 262 | 0.5155 | 0.4580 | 0.5761 |
| 263 | 0.5540 | 0.4568 | 0.5748 |
| 264 | 0.5318 | 0.4580 | 0.5761 |

(d) Virginia Tech Shooting in adjacent states

Table S5: Out-of-sample forecasts for the first $10$ months after the intervention using SRB data grouped into $7$-day periods and fitted with seasonal ARIMA models. Any period of which the observed SRB is outside of the $95\%$ confidence level is marked by an asterisk (*).

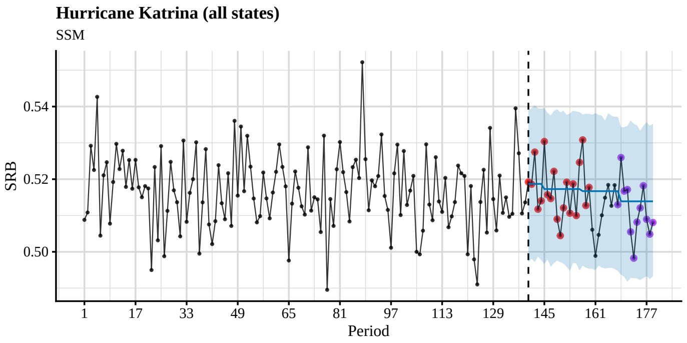


(a) Hurricane Katrina, all states

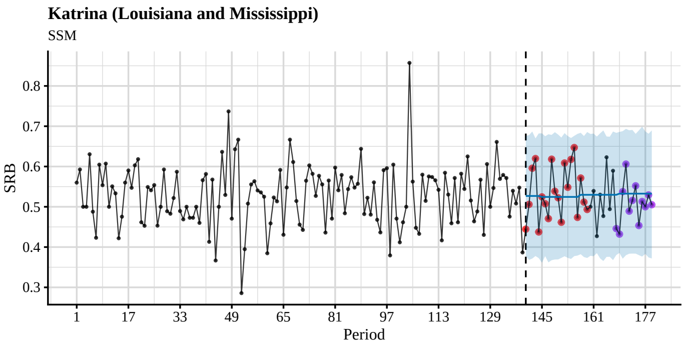


(b) Hurricane Katrina, Louisiana and Mississippi only

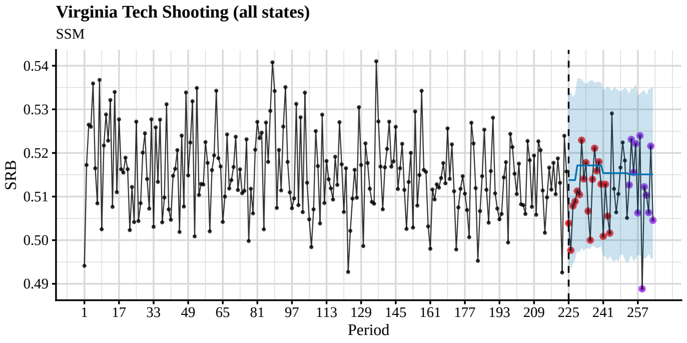


(c) Virginia Tech shooting, all states

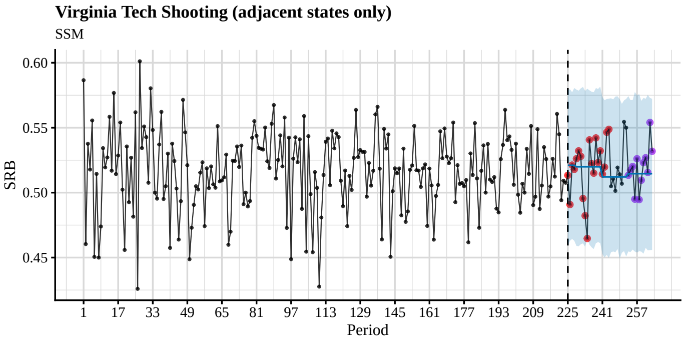


(d) Virginia Tech shooting, adjacent states only

Figure S6: Time series plots and out-of-sample forecasts for SRB data grouped into 28-day periods and fitted with state-space models. The blue shade is the 95% confidence level. The observed SRBs for the first 5 months after the intervention are presented by red dots, whereas the observed SRBs for 7–9 months after the intervention are presented by purple dots. See also Table S6.

| Period | SRB | Lower 95% | Upper 95% |
|---|---|---|---|
| 140 | 0.5192 | 0.4984 | 0.5388 |
| 141 | 0.5186 | 0.4971 | 0.5400 |
| 142 | 0.5275 | 0.4980 | 0.5384 |
| 143 | 0.5117 | 0.4974 | 0.5399 |
| 144 | 0.5140 | 0.4982 | 0.5394 |
| 145 | 0.5304 | 0.4951 | 0.5371 |
| 146 | 0.5157 | 0.4954 | 0.5392 |
| 147 | 0.5147 | 0.4963 | 0.5382 |
| 148 | 0.5222 | 0.4960 | 0.5387 |
| 149 | 0.5090 | 0.4949 | 0.5386 |
| 150 | 0.5045 | 0.4957 | 0.5378 |
| 151 | 0.5121 | 0.4970 | 0.5383 |
| 152 | 0.5192 | 0.4965 | 0.5390 |
| 153 | 0.5106 | 0.4961 | 0.5392 |
| 154 | 0.5187 | 0.4964 | 0.5390 |
| 155 | 0.5100 | 0.4967 | 0.5386 |
| 156 | 0.5246 | 0.4964 | 0.5369 |
| 157 | 0.5308 | 0.4958 | 0.5378 |
| 158 | 0.5128 | 0.4955 | 0.5366 |
| 159 | 0.5177 | 0.4952 | 0.5375 |
| 160 | 0.5061 | 0.4944 | 0.5392 |
| 161 | 0.4989 | 0.4950 | 0.5385 |
| 162 | 0.5047 | 0.4957 | 0.5377 |
| 163 | 0.5101 | 0.4953 | 0.5385 |
| 164 | 0.5149 | 0.4944 | 0.5382 |
| 165 | 0.5184 | 0.4958 | 0.5383 |
| 166 | 0.5127 | 0.4959 | 0.5368 |
| 167 | 0.5184 | 0.4959 | 0.5369 |
| 168 | 0.5130 | 0.4955 | 0.5370 |
| 169 | 0.5260 | 0.4922 | 0.5352 |
| 170 | 0.5167 | 0.4927 | 0.5339 |
| 171 | 0.5171 | 0.4926 | 0.5364 |
| 172 | 0.5055 | 0.4925 | 0.5340 |
| 173 | 0.4983 | 0.4924 | 0.5348 |
| 174 | 0.5082 | 0.4936 | 0.5351 |
| 175 | 0.5121 | 0.4934 | 0.5357 |
| 176 | 0.5182 | 0.4938 | 0.5347 |
| 177 | 0.5089 | 0.4917 | 0.5347 |
| 178 | 0.5049 | 0.4941 | 0.5351 |
| 179 | 0.5081 | 0.4928 | 0.5351 |

(a) Hurricane Katrina in all states

| Period | SRB | Lower 95% | Upper 95% |
|---|---|---|---|
| 140 | 0.4444 | 0.3793 | 0.6830 |
| 141 | 0.5060 | 0.3685 | 0.6803 |
| 142 | 0.5957 | 0.3726 | 0.6806 |
| 143 | 0.6197 | 0.3769 | 0.6859 |
| 144 | 0.4375 | 0.3680 | 0.6870 |
| 145 | 0.5246 | 0.3642 | 0.6749 |
| 146 | 0.5077 | 0.3789 | 0.6835 |
| 147 | 0.4706 | 0.3716 | 0.6780 |
| 148 | 0.6182 | 0.3691 | 0.6732 |
| 149 | 0.5385 | 0.3728 | 0.6870 |
| 150 | 0.5224 | 0.3742 | 0.6746 |
| 151 | 0.4615 | 0.3784 | 0.6823 |
| 152 | 0.6087 | 0.3665 | 0.6824 |
| 153 | 0.5484 | 0.3744 | 0.6809 |
| 154 | 0.6176 | 0.3691 | 0.6777 |
| 155 | 0.6471 | 0.3808 | 0.6810 |
| 156 | 0.4737 | 0.3655 | 0.6821 |
| 157 | 0.5714 | 0.3753 | 0.6799 |
| 158 | 0.5116 | 0.3833 | 0.6857 |
| 159 | 0.4933 | 0.3718 | 0.6812 |
| 160 | 0.5000 | 0.3801 | 0.6791 |
| 161 | 0.5393 | 0.3729 | 0.6883 |
| 162 | 0.4271 | 0.3790 | 0.6797 |
| 163 | 0.5301 | 0.3742 | 0.6746 |
| 164 | 0.4769 | 0.3739 | 0.6827 |
| 165 | 0.6230 | 0.3781 | 0.6797 |
| 166 | 0.4941 | 0.3730 | 0.6901 |
| 167 | 0.5895 | 0.3765 | 0.6865 |
| 168 | 0.4458 | 0.3765 | 0.6815 |
| 169 | 0.4321 | 0.3759 | 0.6864 |
| 170 | 0.5376 | 0.3806 | 0.6869 |
| 171 | 0.6061 | 0.3742 | 0.6965 |
| 172 | 0.4891 | 0.3844 | 0.6839 |
| 173 | 0.5158 | 0.3761 | 0.6805 |
| 174 | 0.5521 | 0.3795 | 0.6878 |
| 175 | 0.4535 | 0.3740 | 0.6792 |
| 176 | 0.5132 | 0.3797 | 0.6959 |
| 177 | 0.5000 | 0.3704 | 0.6787 |
| 178 | 0.5294 | 0.3742 | 0.6895 |
| 179 | 0.5052 | 0.3720 | 0.6802 |

(b) Hurricane Katrina in Louisiana and Mississippi

| Period | SRB | Lower 95% | Upper 95% |
|---|---|---|---|
| 225 | 0.5039 | 0.4926 | 0.5310 |
| 226 | 0.4977 | 0.4939 | 0.5313 |
| 227 | 0.5079 | 0.4941 | 0.5305 |
| 228 | 0.5089 | 0.4934 | 0.5306 |
| 229 | 0.5113 | 0.4954 | 0.5342 |
| 230 | 0.5104 | 0.4964 | 0.5354 |
| 231 | 0.5229 | 0.4972 | 0.5349 |
| 232 | 0.5141 | 0.4953 | 0.5337 |
| 233 | 0.5178 | 0.4954 | 0.5355 |
| 234 | 0.5067 | 0.4977 | 0.5344 |
| 235 | 0.5000 | 0.4966 | 0.5355 |
| 236 | 0.5140 | 0.4955 | 0.5351 |
| 237 | 0.5211 | 0.4967 | 0.5349 |
| 238 | 0.5159 | 0.4958 | 0.5350 |
| 239 | 0.5180 | 0.4970 | 0.5354 |
| 240 | 0.5129 | 0.4963 | 0.5352 |
| 241 | 0.5009 | 0.4943 | 0.5331 |
| 242 | 0.5128 | 0.4945 | 0.5323 |
| 243 | 0.5055 | 0.4938 | 0.5332 |
| 244 | 0.5016 | 0.4948 | 0.5322 |
| 245 | 0.5291 | 0.4940 | 0.5333 |
| 246 | 0.5118 | 0.4947 | 0.5342 |
| 247 | 0.5064 | 0.4946 | 0.5331 |
| 248 | 0.5106 | 0.4952 | 0.5328 |
| 249 | 0.5167 | 0.4944 | 0.5332 |
| 250 | 0.5224 | 0.4940 | 0.5335 |
| 251 | 0.5183 | 0.4948 | 0.5322 |
| 252 | 0.5051 | 0.4942 | 0.5333 |
| 253 | 0.5127 | 0.4948 | 0.5324 |
| 254 | 0.5231 | 0.4936 | 0.5325 |
| 255 | 0.5155 | 0.4936 | 0.5325 |
| 256 | 0.5221 | 0.4947 | 0.5335 |
| 257 | 0.5063 | 0.4938 | 0.5341 |
| 258 | 0.5240 | 0.4947 | 0.5327 |
| 259(*) | 0.4888 | 0.4944 | 0.5327 |
| 260 | 0.5122 | 0.4946 | 0.5337 |
| 261 | 0.5103 | 0.4944 | 0.5326 |
| 262 | 0.5063 | 0.4936 | 0.5337 |
| 263 | 0.5216 | 0.4938 | 0.5326 |
| 264 | 0.5046 | 0.4938 | 0.5341 |

(c) Virginia Tech Shooting in all states

| Period | SRB | Lower 95% | Upper 95% |
|---|---|---|---|
| 225 | 0.5134 | 0.4612 | 0.5784 |
| 226 | 0.4908 | 0.4613 | 0.5839 |
| 227 | 0.5212 | 0.4622 | 0.5824 |
| 228 | 0.5179 | 0.4640 | 0.5814 |
| 229 | 0.5261 | 0.4644 | 0.5794 |
| 230 | 0.5322 | 0.4627 | 0.5820 |
| 231 | 0.5280 | 0.4604 | 0.5799 |
| 232 | 0.4955 | 0.4590 | 0.5822 |
| 233 | 0.4822 | 0.4609 | 0.5793 |
| 234 | 0.4647 | 0.4554 | 0.5789 |
| 235 | 0.5406 | 0.4622 | 0.5778 |
| 236 | 0.5223 | 0.4625 | 0.5801 |
| 237 | 0.5150 | 0.4609 | 0.5819 |
| 238 | 0.5421 | 0.4593 | 0.5778 |
| 239 | 0.5231 | 0.4603 | 0.5750 |
| 240 | 0.5322 | 0.4610 | 0.5764 |
| 241 | 0.5143 | 0.4519 | 0.5705 |
| 242 | 0.5197 | 0.4484 | 0.5681 |
| 243 | 0.5465 | 0.4514 | 0.5725 |
| 244 | 0.5487 | 0.4530 | 0.5713 |
| 245 | 0.5049 | 0.4547 | 0.5747 |
| 246 | 0.5105 | 0.4553 | 0.5755 |
| 247 | 0.5014 | 0.4525 | 0.5729 |
| 248 | 0.5193 | 0.4528 | 0.5704 |
| 249 | 0.5141 | 0.4513 | 0.5735 |
| 250 | 0.5068 | 0.4526 | 0.5735 |
| 251 | 0.5545 | 0.4548 | 0.5722 |
| 252 | 0.5500 | 0.4525 | 0.5697 |
| 253 | 0.5133 | 0.4515 | 0.5723 |
| 254 | 0.5172 | 0.4514 | 0.5741 |
| 255 | 0.5201 | 0.4574 | 0.5718 |
| 256 | 0.4949 | 0.4575 | 0.5749 |
| 257 | 0.5261 | 0.4548 | 0.5744 |
| 258 | 0.4946 | 0.4553 | 0.5765 |
| 259 | 0.5096 | 0.4553 | 0.5750 |
| 260 | 0.5231 | 0.4550 | 0.5745 |
| 261 | 0.5269 | 0.4571 | 0.5737 |
| 262 | 0.5155 | 0.4568 | 0.5762 |
| 263 | 0.5540 | 0.4540 | 0.5755 |
| 264 | 0.5318 | 0.4576 | 0.5720 |

(d) Virginia Tech Shooting in adjacent states

Table S6: Out-of-sample forecasts for the first 10 months after the intervention using SRB data grouped into 7-day periods and fitted with state-space models. Any period of which the observed SRB is outside of the 95% confidence level is marked by an asterisk (*).

### V Swedish SRB and Meteorological Observations

Using the data downloaded from the World Bank (https://climateknowledgeportal.worldbank.org/download-data), we performed a Pearson's correlation test and a Granger causality test. The $p$-values for the null hypotheses of the nonexistence of correlation (Student's $t$-test) and Granger causality ($F$-test) are listed in Table S7. We could not establish associations between the SRB and either of the meteorological observations between years 1991 and 2013.

| | Temperature | Precipitation |
|---|---|---|
| $t$-test | 0.156 | 0.765 |
| $F$-test | 0.269 | 0.228 |

Table S7: $p$-values for $t$- and $F$-tests on the correlation between Sweden's SRB and temperature and precipitation in Sweden

In addition, we performed logistic regression using the following:

| Factor | $\Delta$IC | SE |
|---|---|---|
| At risk of poverty | 0.9934 | 3.5615 |
| $SO_2$ | 0.8399 | 3.6321 |
| $NO_2$ | -0.5075 | 3.2862 |
| Proportion foreign nationals | -1.8912 | 2.2786 |
| Black smoke | -2.0551 | 2.7690 |
| P80/P20 | -2.2833 | 2.2427 |
| Car density | -2.6865 | 2.4586 |
| Population density | -2.7654 | 2.1618 |
| Gini | -2.7838 | 2.2333 |
| Mean income | -3.4858 | 2.1006 |
| PAH | -3.7904 | 2.0331 |
| VOC | -4.3064 | 1.8581 |
| $PM_{10}$ | -4.4771 | 1.7049 |
| $PM_{2.5}$ | -4.9524 | 1.4270 |
| Median income | -5.1749 | 1.9721 |

Table S8: Differences in information criteria ($\Delta$IC) and their standard errors (SE) of individual factors at the kommun (municipality) level, with random effect at the län (county) level

Table S9: Differences in information criteria ($\Delta$IC) and their standard errors (SE) of individual factors at the län (county) level

| Factor | $\Delta$IC | SE |
|---|---|---|
| diseases of the respiratory system men | 3.3012 | 2.5949 |
| good health men and women | 1.4435 | 2.4498 |
| diseases of the circulatory system women | 1.1142 | 3.2312 |
| serious motor disabilities women | 0.9167 | 3.3112 |
| smoke and or take snuff daily men | 0.8522 | 2.1592 |
| unmet need for medical care men | 0.7556 | 2.4153 |
| motor disabilities men | 0.6604 | 3.1667 |
| high blood pressure women | 0.2627 | 3.0594 |
| smoke and or take snuff daily women | 0.2378 | 3.1951 |
| impaired hearing men | 0.0979 | 2.7779 |
| diabetes men | 0.0969 | 3.1908 |
| diseases of the circulatory system men | 0.0838 | 2.7351 |
| diseases of the respiratory system men and women | -0.1382 | 2.9142 |
| serious pain total men and women | -0.1481 | 2.3953 |
| serious motor disabilities men and women | -0.1740 | 3.0316 |
| serious problems of anxiety worry fear men and women | -0.2266 | 3.5719 |
| unmet need for medical care women | -0.2308 | 2.5992 |
| poor health women | -0.3233 | 2.2780 |
| smoke daily women | -0.3289 | 3.2831 |
| impaired hearing women | -0.5162 | 2.7654 |
| problems of anxiety worry fear men | -0.5694 | 2.2561 |
| impaired vision men and women | -0.7338 | 2.4952 |
| obese BMI 30 or more men | -1.0034 | 3.1003 |
| serious motor disabilities men | -1.0759 | 2.2428 |
| serious pain in shoulders neck women | -1.1545 | 2.2927 |
| impaired vision women | -1.1893 | 2.4814 |
| serious pain in hands elbows or knees men | -1.2649 | 3.0558 |
| smoke daily men | -1.5168 | 2.7761 |
| diseases of the musculoskeletal system and connective tissue men | -1.5226 | 2.4445 |
| diseases of the skin men and women | -1.6969 | 3.6015 |
| diseases of the musculoskeletal system and connective tissue men and women | -1.7217 | 2.6642 |
| diabetes women | -1.8868 | 2.4746 |
| severe problems from a long term illness women | -2.1122 | 2.7674 |
| serious pain in shoulders neck men | -2.1671 | 2.8550 |
| trouble sleeping men and women | -2.1719 | 2.1819 |
| endocrine diseases men and women | -2.1860 | 2.4510 |
| trouble sleeping women | -2.1972 | 3.5518 |
| endocrine diseases women | -2.3205 | 2.5285 |
| dentist appointments during a 12 month period men and women | -2.3418 | 2.8350 |
| serious pain total men | -2.3436 | 4.0538 |
| problems of anxiety worry fear men and women | -2.4328 | 2.5539 |
| poor health men | -2.4561 | 2.3493 |
| serious pain in back or hips men and women | -2.6307 | 3.2649 |

Continued on next page

Table S9: Differences in information criteria (ΔIC) and their standard errors (SE) of individual factors at the län (county) level

| Factor | ΔIC | SE |
|---|---|---|
| diseases of the circulatory system men and women | -2.7013 | 2.3490 |
| high blood pressure men and women | -2.7124 | 2.1525 |
| overweight or obese BMI 25 or more men and women | -2.7407 | 2.4722 |
| serious pain total women | -2.7984 | 2.2393 |
| diseases of the musculoskeletal system and connective tissue women | -2.8772 | 2.9321 |
| diseases of the skin men | -3.0005 | 2.7985 |
| dentist appointments during a 12 month period men | -3.0185 | 2.9935 |
| unmet need for medical care men and women | -3.0341 | 1.7253 |
| serious pain in hands, elbows, or knees women | -3.1847 | 3.1895 |
| diabetes men and women | -3.2320 | 2.2913 |
| serious problems of anxiety, worry, fear men | -3.2685 | 2.7144 |
| serious pain in shoulders, neck men and women | -3.2852 | 2.7533 |
| doctor appointments during a three month period men | -3.3952 | 1.9437 |
| heart disease men and women | -3.4409 | 2.5030 |
| overweight or obese BMI 25 or more men | -3.4821 | 2.5026 |
| motor disabilities women | -3.5407 | 3.4502 |
| obese BMI 30 or more men and women | -3.6252 | 2.4101 |
| doctor appointments during a three month period men and women | -3.6527 | 1.6407 |
| endocrine diseases men | -3.7796 | 2.4763 |
| overweight or obese BMI 25 0 or more women | -3.9447 | 2.8735 |
| severe problems from a long term illness men and women | -3.9898 | 2.9988 |
| good health women | -4.0257 | 2.8712 |
| heart disease men | -4.1542 | 2.1345 |
| poor health men and women | -4.3023 | 3.0782 |
| take snuff daily men and women | -4.3065 | 3.0176 |
| problems of anxiety worry fear women | -4.3155 | 2.2453 |
| serious pain in hands elbows or knees men and women | -4.3937 | 2.3697 |
| impaired hearing men and women | -4.5371 | 2.0341 |
| diseases of the digestive system men and women | -4.6030 | 2.6724 |
| serious pain in back or hips women | -4.6210 | 2.2235 |
| severe problems from a long term illness men | -4.7071 | 2.3077 |
| dentist appointments during a 12 month period women | -4.7839 | 2.6277 |
| take snuff daily men | -4.9910 | 2.1400 |
| impaired vision men | -5.1140 | 2.2114 |
| unmet need for dental care men | -5.6746 | 2.5992 |
| diseases of the respiratory system women | -5.7384 | 2.5305 |
| high blood pressure men | -5.7527 | 3.8451 |
| serious pain in back or hips men | -5.8251 | 3.5053 |
| diseases of the digestive system men | -5.9368 | 2.6694 |
| unmet need for dental care men and women | -6.1628 | 1.9368 |
| heart disease women | -6.2097 | 1.9162 |
| take snuff daily women | -6.3476 | 2.0574 |
| obese BMI 30 0 or more women | -6.4418 | 2.0136 |
| motor disabilities men and women | -6.7537 | 1.9335 |

Continued on next page

Table S9: Differences in information criteria ($\Delta$IC) and their standard errors (SE) of individual factors at the län (county) level

| Factor | $\Delta$IC | SE |
|---|---|---|
| smoke and or take snuff daily men and women | -6.7639 | 2.4234 |
| trouble sleeping men | -6.7890 | 3.4973 |
| smoke daily men and women | -6.8021 | 4.0868 |
| diseases of the nervous system and the sensory organs men | -6.8766 | 1.8000 |
| diseases of the skin women | -7.0391 | 2.9732 |
| diseases of the nervous system and the sensory organs men and women | -7.0903 | 2.1524 |
| diseases of the digestive system women | -7.0938 | 2.0396 |
| doctor appointments during a three month period women | -7.1649 | 4.1678 |
| diseases of the nervous system and the sensory organs women | -7.1994 | 2.4814 |
| good health men | -7.4083 | 1.7240 |
| unmet need for dental care women | -8.1221 | 3.4038 |
| serious problems of anxiety worry fear women | -8.7865 | 1.7444 |

### VI Contingency Table Analysis

Table S10 is the full contingency table for testing the association between physical injury, infections, and neuropsychiatric disorders (stratified by before/during diagnosis) and SRB.

| Phys_1y | Infection_1y | Neuropsych_1y | Phys_older | Infection_older | Neuropsych_older | F | M | Total |
|---|---|---|---|---|---|---|---|---|
| 0 | 0 | 0 | 0 | 0 | 0 | 693079 | 733474 | 1426553 |
| 0 | 0 | 0 | 0 | 0 | 1 | 16835 | 17539 | 34374 |
| 0 | 0 | 0 | 0 | 1 | 0 | 65169 | 68766 | 133935 |
| 0 | 0 | 0 | 0 | 1 | 1 | 15297 | 16215 | 31512 |
| 0 | 0 | 0 | 1 | 0 | 0 | 5700 | 5945 | 11645 |
| 0 | 0 | 0 | 1 | 0 | 1 | 1265 | 1420 | 2685 |
| 0 | 0 | 0 | 1 | 1 | 0 | 5031 | 5250 | 10281 |
| 0 | 0 | 0 | 1 | 1 | 1 | 2424 | 2565 | 4989 |
| 0 | 0 | 1 | 0 | 0 | 0 | 30176 | 31833 | 62009 |
| 0 | 0 | 1 | 0 | 0 | 1 | 10905 | 11480 | 22385 |
| 0 | 0 | 1 | 0 | 1 | 0 | 5456 | 5688 | 11144 |
| 0 | 0 | 1 | 0 | 1 | 1 | 8629 | 9092 | 17721 |
| 0 | 0 | 1 | 1 | 0 | 0 | 464 | 537 | 1001 |
| 0 | 0 | 1 | 1 | 0 | 1 | 808 | 857 | 1665 |
| 0 | 0 | 1 | 1 | 1 | 0 | 510 | 477 | 987 |
| 0 | 0 | 1 | 1 | 1 | 1 | 1488 | 1554 | 3042 |
| 0 | 1 | 0 | 0 | 0 | 0 | 65857 | 69326 | 135183 |
| 0 | 1 | 0 | 0 | 0 | 1 | 3926 | 4229 | 8155 |
| 0 | 1 | 0 | 0 | 1 | 0 | 24502 | 25721 | 50223 |
| 0 | 1 | 0 | 0 | 1 | 1 | 6555 | 6952 | 13507 |
| 0 | 1 | 0 | 1 | 0 | 0 | 1230 | 1241 | 2471 |
| 0 | 1 | 0 | 1 | 0 | 1 | 352 | 385 | 737 |
| 0 | 1 | 0 | 1 | 1 | 0 | 1962 | 1993 | 3955 |
| 0 | 1 | 0 | 1 | 1 | 1 | 1077 | 1185 | 2262 |
| 0 | 1 | 1 | 0 | 0 | 0 | 9609 | 10417 | 20026 |
| 0 | 1 | 1 | 0 | 0 | 1 | 3666 | 3960 | 7626 |
| 0 | 1 | 1 | 0 | 1 | 0 | 3301 | 3422 | 6723 |
| 0 | 1 | 1 | 0 | 1 | 1 | 5243 | 5653 | 10896 |
| 0 | 1 | 1 | 1 | 0 | 0 | 174 | 173 | 347 |
| 0 | 1 | 1 | 1 | 0 | 1 | 312 | 316 | 628 |
| 0 | 1 | 1 | 1 | 1 | 0 | 271 | 313 | 584 |
| 0 | 1 | 1 | 1 | 1 | 1 | 977 | 1006 | 1983 |
| 1 | 0 | 0 | 0 | 0 | 0 | 5894 | 6180 | 12074 |
| 1 | 0 | 0 | 0 | 0 | 1 | 434 | 420 | 854 |
| 1 | 0 | 0 | 0 | 1 | 0 | 1484 | 1565 | 3049 |
| 1 | 0 | 0 | 0 | 1 | 1 | 426 | 469 | 895 |
| 1 | 0 | 0 | 1 | 0 | 0 | 245 | 245 | 490 |
| 1 | 0 | 0 | 1 | 0 | 1 | 69 | 70 | 139 |
| 1 | 0 | 0 | 1 | 1 | 0 | 227 | 221 | 448 |
| 1 | 0 | 0 | 1 | 1 | 1 | 117 | 138 | 255 |
| 1 | 0 | 1 | 0 | 0 | 0 | 1329 | 1454 | 2783 |
| 1 | 0 | 1 | 0 | 0 | 1 | 527 | 550 | 1077 |
| 1 | 0 | 1 | 0 | 1 | 0 | 258 | 286 | 544 |
| 1 | 0 | 1 | 0 | 1 | 1 | 436 | 482 | 918 |
| 1 | 0 | 1 | 1 | 0 | 0 | 28 | 50 | 78 |
| 1 | 0 | 1 | 1 | 0 | 1 | 195 | 203 | 398 |
| 1 | 0 | 1 | 1 | 1 | 0 | 28 | 43 | 71 |
| 1 | 0 | 1 | 1 | 1 | 1 | 235 | 256 | 491 |
| 1 | 1 | 0 | 0 | 0 | 0 | 2122 | 2204 | 4326 |
| 1 | 1 | 0 | 0 | 0 | 1 | 181 | 182 | 363 |
| 1 | 1 | 0 | 0 | 1 | 0 | 929 | 903 | 1832 |
| 1 | 1 | 0 | 0 | 1 | 1 | 288 | 305 | 593 |
| 1 | 1 | 0 | 1 | 0 | 0 | 69 | 79 | 148 |
| 1 | 1 | 0 | 1 | 0 | 1 | 27 | 31 | 58 |
| 1 | 1 | 0 | 1 | 1 | 0 | 143 | 134 | 277 |
| 1 | 1 | 0 | 1 | 1 | 1 | 81 | 87 | 168 |
| 1 | 1 | 1 | 0 | 0 | 0 | 650 | 713 | 1363 |
| 1 | 1 | 1 | 0 | 0 | 1 | 249 | 263 | 512 |
| 1 | 1 | 1 | 0 | 1 | 0 | 214 | 226 | 440 |
| 1 | 1 | 1 | 0 | 1 | 1 | 408 | 427 | 835 |
| 1 | 1 | 1 | 1 | 0 | 0 | 22 | 15 | 37 |
| 1 | 1 | 1 | 1 | 0 | 1 | 66 | 84 | 150 |
| 1 | 1 | 1 | 1 | 1 | 0 | 35 | 46 | 81 |
| 1 | 1 | 1 | 1 | 1 | 1 | 204 | 203 | 407 |
| Total | | | | | | 1009870 | 1067518 | 2077388 |

Table S10: Contingency table of maternal diagnosis history versus the sex of livebirths

### VII Dictionary of factors and their definitions

Table S11: List of variable names used in the main text and their corresponding definitions and units (if applicable)

| Variable Name | Variable Definition | Units |
|---|---|---|
| A_PM10_mean_ln | particulate matter under ten micrometers in aerodynamic diameter (PM10) | ln-$\mu$g/m3 |
| A_PM25_mean | particulate matter under 2.5 micrometers in aerodynamic diameter (PM2.5) | ln-$\mu$g/m3 |
| A_NO2_mean_ln | nitrogen dioxide (NO2) | ln-ppb |
| A_SO2_mean_ln | sulfur dioxide (SO2) | ln-ppb |
| A_O3_mean_ln | ozone (O3) | ln-ppm |
| A_CO_mean_ln | carbon monoxide (CO) | ln-ppm |
| A_TeCA_ln | 1,1,2,2-tetrachloroethane | ln-tons |
| A_112TCA_ln | 1,1,2-trichloroethane | ln-tons |
| A_DBCP_ln | 1,2-dibromo-3-chloropropane | ln-tons |
| A_TDI_ln | 2,4-toluene diisocyanate | ln-tons |
| A_2Clacephen_ln | 2-chloroacetophenone | ln-tons |
| A_2NP_ln | 2-nitropropane | ln-tons |
| A_PNP_ln | 4-nitrophenol | ln-tons |
| A_CH3CN_ln | acetonitrile | ln-tons |
| A_Acetophenone_ln | acetophenone | ln-tons |
| A_Acrolein_ln | acrolein | ln-tons |
| A_Acrylic_acid_ln | acrylic acid | ln-tons |
| A_C3H3N_ln | acrylonitrile | ln-tons |
| A_Sb_ln | antimony compounds | ln-tons |
| A_Benzidine_ln | benzidine | ln-tons |
| A_Benzyl_Cl_ln | benzyl chloride | ln-tons |
| A_Be_ln | beryllium compounds | ln-tons |
| A_biphenyl_ln | biphenyl | ln-tons |
| A_DEHP_ln | bis-2-ethylhexyl phthalate | ln-tons |
| A_Bromoform_ln | bromoform | ln-tons |
| A_Cd_ln | cadmium compounds | ln-tons |
| A_CS2_ln | carbon disulfide | ln-tons |
| A_CCl4 | carbon tetrachloride | tons |
| A_CS_ln | carbon sulfide | ln-tons |
| A_Cl_ln | chlorine | ln-tons |
| A_C6H5Cl_ln | chlorobenzene | ln-tons |
| A_chloroform_ln | chloroform | ln-tons |
| A_Chloroprene_ln | chloroprene | ln-tons |
| A_Cr_ln | chromium compounds | ln-tons |
| A_Cresol_ln | cresol/cresylic acid | ln-tons |
| A_Cumene_ln | cumene | ln-tons |
| A_CN_ln | cyanide compounds | ln-tons |
| A_DBP_ln | dibutylphthalate | ln-tons |
| A_Diesel_ln | diesel engine emissions | ln-tons |
| A_DMF_ln | dimethyl formamide | ln-tons |
| A_Me2_phthalate_ln | dimethyl phthalates | ln-tons |
| A_Me2SO4_ln | dimethyl sulfate | ln-tons |
| A_ECH_ln | epichlorohydrin | ln-tons |
| A_Etacrylate_ln | ethyl acrylate | ln-tons |
| A_EtCl_ln | ethyl chloride | ln-tons |
| A_EDB_ln | ethylene dibromide | ln-tons |
| A_EDC_ln | ethylene dichloride | ln-tons |
| A_EGLY_ln | ethylene glycol | ln-tons |
| A_EOx_ln | ethylene oxide | ln-tons |
| A_EdCl2_ln | ethylidene dichloride | ln-tons |
| A_Glycol_ethers_ln | glycol ethers | ln-tons |
| A_HCB_ln | hexachlorobenzene | ln-tons |
| A_HCBD_ln | hexachlorobutadiene | ln-tons |
| A_HCCPD_ln | hexachlorocyclopentadiene | ln-tons |
| A_Hexane_ln | hexane | ln-tons |
| A_N2H2_ln | hydrazine | ln-tons |
| A_HCl_ln | hydrochloric acid | ln-tons |
| A_Isophorone_ln | isophorone | ln-tons |
| A_Pb_ln | lead compounds | ln-tons |
| A_Mn_ln | manganese compounds | ln-tons |
| A_Hg_ln | mercury compounds | ln-tons |
| A_MeOH_ln | methanol | ln-tons |
| A_MIBK_ln | methyl isobutyl ketone | ln-tons |
| A_MMA_ln | methyl methacrylate | ln-tons |
| A_MeCl_ln | methyl chloride | ln-tons |
| A_Mehydrazine_ln | methylhydrazine | ln-tons |
| A_MTBE_ln | MTBE | ln-tons |
| A_nitrobenzene_ln | nitrobenzene | ln-tons |
| A_DMA_ln | N,N-dimethylaniline | ln-tons |
| A_otoluidine_ln | o-toluidine | ln-tons |
| A_PAHPOM_ln | PAH/POM | ln-tons |
| A_PCP_ln | pentachlorophenol | ln-tons |
| A_PH3_ln | phosphine | ln-tons |
| A_P_ln | phosphorus | ln-tons |
| A_PCBs_ln | polychlorinated biphenyls | ln-tons |
| A_ProCl2_ln | propylene dichloride | ln-tons |
| A_ProO_ln | propylene oxide | ln-tons |
| A_Quinoline_ln | quinoline | ln-tons |
| A_Se_ln | selenium compounds | ln-tons |
| A_Styrene_ln | styrene | ln-tons |
| A_Cl4C2_ln | tetrachloroethylene | ln-tons |
| A_Toluene_ln | toluene | ln-tons |
| A_C2HCl3_ln | trichloroethylene | ln-tons |
| A_Et3N_ln | triethylamine | ln-tons |
| A_VyAc_ln | vinyl acetate | ln-tons |

Continued on next page

Table S11: List of variable names used in the main text and their corresponding definitions and units (if applicable)

| Variable Name | Variable Definition | Units |
|---|---|---|
| A_VyCl_ln | vinyl chloride | ln-tons |
| A_11DCE_ln | vinylidene chloride | ln-tons |
| D303_Percent | % of stream length impaired in county | percent |
| SEWAGENPDESperKM | sewage permits per 1000 km of stream in county | permits/1000km |
| INDNPDESperKM | industrial permits per 1000 km of stream in county | permits/1000km |
| STORMNPDESperKM | stormwater permits per 1000 km of stream in county | permits/1000km |
| numDays_Close_Activity_tot | # of days closed per event in county 2000-2005 | days |
| numDays_Cont_Activity_tot | # of days per contamination advisory event in county 2000-2005 | days |
| numDays_Rain_Activity_tot | # of days per rain advisory event in county 2000-2005 | days |
| Per_TotPopSS_ave | percent of population on self supply, average 2000&2005 | percent |
| Per_PSWithSW_ave | percent of public supply population which is on surface water, average 2000&2005 | percent |
| Ca_ln_ave | calcium (Ca) precipitation weighted mean | ln mg/L |
| Mg_ln_ave | magnesium (Mg) precipitation weighted mean | ln mg/L |
| K_ln_ave | potassium (K) precipitation weighted mean | ln mg/L |
| Na_ln_ave | sodium (Na) precipitation weighted mean | ln mg/L |
| NH4_mean_ave | ammonium (NH4) precipitation weighted mean | mg/L |
| NO3_mean_ave | nitrate (NO3) precipitation weighted mean | mg/L |
| Cl_ln_ave | chloride (Cl) precipitation weighted mean | ln mg/L |
| SO4_mean_ave | sulfate (SO4) precipitation weighted mean | mg/L |
| Hg_ln_ave | total mercury (Hg) deposition | ln mg/L |
| AvgOfD3_ave | % of county drought – extreme (3-D4) | percent |
| W_As_ln | arsenic | ln mg/L |
| W_Ba_ln | barium | ln mg/L |
| W_Cd_ln | cadmium | ln mg/L |
| W_Cr_ln | chromium | ln mg/L |
| W_CN_ln | cyanide | ln mg/L |
| W_FL_ln | fluoride | ln mg/L |
| W_HG_ln | mercury (inorganic) | ln mg/L |
| W_NO3_ln | nitrate | ln mg/L |
| W_NO2_ln | nitrite | ln mg/L |
| W_SE_ln | selenium | ln mg/L |
| W_Sb_ln | antimony | ln mg/L |
| W_Be_ln | beryllium | ln mg/L |
| W_Tl_ln | thallium | ln mg/L |
| W_Endrin_ln | endrin | ln mg/L |
| W_Lindane_ln | lindane | ln mg/L |
| W_methoxychlor_ln | methoxychlor | $\ln \mu g/L$ |
| W_Toxaphene_ln | toxaphene | $\ln \mu g/L$ |
| W_Dalapon_ln | dalapon | $\ln \mu g/L$ |
| W_DEHA_ln | di(2-ethylhexyl)adipate (DEHA) | $\ln \mu g/L$ |
| W_Oxamyl_ln | oxamyl (Vydate) | $\ln \mu g/L$ |
| W_Simazine_ln | simazine | $\ln \mu g/L$ |
| W_DEHP_ln | Di(2-ethylhexyl) phthalate (DEHP) | $\ln \mu g/L$ |
| W_Picloram_ln | picloram | $\ln \mu g/L$ |
| W_Dinoseb_ln | dinoseb | $\ln \mu g/L$ |
| W_HCCPD_ln | hexachlorocyclopentadiene | $\ln \mu g/L$ |
| W_Carbofuran_ln | carbofuran | $\ln \mu g/L$ |
| W_atrazine_ln | atrazine | $\ln \mu g/L$ |
| W_Alachlor_ln | alachlor | $\ln \mu g/L$ |
| W_Heptachlor_ln | heptachlor | $\ln \mu g/L$ |
| W_Heptachlor_epox_ln | heptachlor epoxide | $\ln \mu g/L$ |
| W_24D_ln | 2,4-D (2,4-Dichlorophenoxyacetic acid) | $\ln \mu g/L$ |
| W_HCB_ln | hexachlorobenzene | $\ln \mu g/L$ |
| W_BenzoAP_ln | benzo[a]pyrene | $\ln \mu g/L$ |
| W_PCP_ln | pentachlorophenol | $\ln \mu g/L$ |
| W_124TCIB_ln | 1,2,4-trichlorobenzene | $\ln \mu g/L$ |
| W_PCB_ln | polychlorinated biphenyls (PCBs) | $\ln \mu g/L$ |
| W_DBCP_ln | 1,2-dibromo-3-chloropropane (DBCP) | $\ln \mu g/L$ |
| W_EBD_ln | ethylene dibromide (EDB) | $\ln \mu g/L$ |
| W_xylenes_ln | xylenes | $\ln \mu g/L$ |
| W_Chlordane_ln | chlordane | $\ln \mu g/L$ |
| W_DCM_ln | dichloromethane (methylene chloride) | $\ln \mu g/L$ |
| W_ODCB_ln | 1,2-dichlorobenzene (o-dichlorobenzene) | $\ln \mu g/L$ |
| W_PDCB_ln | 1,4-dichlorobenzene (p-dichlorobenzene) | $\ln \mu g/L$ |
| W_VCM_ln | vinyl chloride | $\ln \mu g/L$ |
| W_11DCE_ln | 1,1-dichloroethylene | $\ln \mu g/L$ |
| W_t12DCE_ln | trans-1,2-Dichloroethylene | $\ln \mu g/L$ |
| W_EDC_ln | 1,2-dichloroethane (Ethylene Dichloride) | $\ln \mu g/L$ |
| W_111trichlorane_ln | 1,1,1-trichloroethane | $\ln \mu g/L$ |
| W_CCl4_ln | carbon tetrachloride | $\ln \mu g/L$ |
| W_PDC_ln | 1,2-dichloropropane | $\ln \mu g/L$ |
| W_Trichlorene_ln | trichloroethylene | $\ln \mu g/L$ |
| W_112TCA_ln | 1,1,2-trichloroethane | $\ln \mu g/L$ |
| W_C2Cl4_ln | tetrachloroethylene | $\ln \mu g/L$ |
| W_benzene_ln | benzene | $\ln \mu g/L$ |
| W_Cl1benz_ln | monochlorobenzene (chlorobenzene) | $\ln \mu g/L$ |
| W_Toluene_ln | toluene | $\ln \mu g/L$ |
| W_ethylbenz_ln | ethylbenzene | $\ln \mu g/L$ |
| W_styrene_ln | styrene | $\ln \mu g/L$ |
| W_DCE_ln | alpha particles | $\ln \mu g/L$ |
| W_alpha | cis-1,2-dichloroethylene | PCl/L |
| W_SILVEX_ln | silvex | $\ln \mu g/L$ |
| pct_harvest_acres | harvested acreage | percent |
| pct_irrigated_acres_ln | irrigated acreage | ln-percent |
| farms_per_acre_ln | farms per acre | ln-(number farms/total acres) |
| pct_manure_acres_ln | manure applied | ln-percent |
| pct_nematode_acres_ln | chemicals used to control nematodes | ln-percent |
| pct_disease_acres_ln | chemicals used to control disease | ln-percent |
| pct_defoliate_acres_ln | chemicals used to defoliate/control growth/thin fruit | ln-percent |
| pct_au_ln | animal units | ln-percent |

Continued on next page

Table S11: List of variable names used in the main text and their corresponding definitions and units (if applicable)

| Variable Name | Variable Definition | Units |
|---|---|---|
| herbicides_ln | herbicides | ln-pounds |
| fungicides_ln | fungicides | ln-pounds |
| insecticides_ln | insecticides | ln-pounds |
| mean_as_ln | arsenic | ln-ppm |
| mean_se_ln | selenium | ln-ppm |
| mean_hg_ln | mercury | ln-ppm |
| mean_pb_ln | lead | ln-ppm |
| mean_zn_ln | zinc | ln-ppm |
| mean_cu_ln | copper | ln-ppm |
| mean_na__pct_ln | sodium | ln-weighted percent |
| mean_mg_pct_ln | magnesium | ln-weighted percent |
| mean_ti_pct_ln | titanium | ln-weighted percent |
| mean_ca_pct_ln | calcium | ln-weighted percent |
| mean_fe_pct_ln | iron | ln-weighted percent |
| mean_al_pct | aluminum | weighted percent |
| mean_p_pct | phosphorus | weighted percent |
| facilities_rate_log | facilities per county pop | ln-rate |
| radon_zone | radon zone | radon category |
| HWYPROP | proportion of roads that are highway | miles highways / miles total roads |
| RYPROP | proportion of roads that are primary streets | miles primary streets / miles total roads |
| fatal_rate_log | traffic fatality rate | ln-rate |
| pct_pub_transport_log | percent of population using public transport | ln-percent |
| rate_al_pn_gm_env_log | vice-related businesses | ln-rate |
| rate_ent_env_log | entertainment-related businesses | ln-rate |
| rate_ed_env_log | education-related businesses | ln-rate |
| rate_food_env_neg | negative food related businesses | rate |
| rate_food_env_pos_log | positive food related businesses | ln-rate |
| rate_hc_env_log | health care-related businesses | ln-rate |
| rate_rec_env_log | recreation-related businesses | ln-rate |
| rate_trans_env_log | transportation-related businesses | ln-rate |
| rate_civic_env_log | civic-related businesses | ln-rate |
| to_unit_rate_log | total subsidized housing units | ln-rate |
| pct_rent_occ | percent renter occupied | count county occupied rental units / total county units |
| pct_vac_units | percent vacant units | count county vacant units / total housing units |
| med_hh_value | median household value | dollars |
| med_hh_inc | median household income | dollars |
| pct_pers_lt_pov | percent persons less than poverty level | count county persons below poverty / county population |
| pct_no_eng | percent no English | count county non-English speaking / county population |
| pct_hs_more | percent earning greater than high school education | count county more than high school / county population |
| pct_unemp | percent unemployed | count county unemployed / county population |
| work_out_co | percent work outside county | count county work outside county / county population |
| med_rooms | median number rooms per house | sum county count of rooms / county housing units |
| pct_mt_10units_log | percent of housing with more than 10 units | ln-percent |
| violent_rate_log | mean number of violent crimes per capita | ln-rate |
| fips | FIPS code to state and county level | N/A |
| county_name | name of county | N/A |
| state | state | N/A |
| cat_rucc | rural-urban continuum code category | N/A |
| air_EQI_22July2013 | non-stratified air domain index | N/A |
| water_EQI_22July2013 | non-stratified water domain index | N/A |
| land_EQI_22July2013 | non-stratified land domain index | N/A |
| sociod_EQI_22July2013 | non-stratified sociodemographic domain index | N/A |
| built_EQI_22July2013 | non-stratified built environment domain index | N/A |
| EQI_22July2013 | non-stratified environmental quality index | N/A |
| RUCC1_air_EQI_22July2013 | metropolitian-urbanized strata air domain index | N/A |
| RUCC1_water_EQI_22July2013 | metropolitian-urbanized strata water domain index | N/A |
| RUCC1_land_EQI_22July2013 | metropolitian-urbanized strata land domain index | N/A |
| RUCC1_sociod_EQI_22July2013 | metropolitian-urbanized strata sociodemographic domain index | N/A |
| RUCC1_built_EQI_22July2013 | metropolitian-urbanized strata built environment domain index | N/A |
| RUCC1_EQI_22July2013 | metropolitian-urbanized strata environmental quality index | N/A |
| RUCC2_air_EQI_22July2013 | non-metropolitian-urbanized strata air domain index | N/A |
| RUCC2_water_EQI_22July2013 | non-metropolitian-urbanized strata water domain index | N/A |
| RUCC2_land_EQI_22July2013 | non-metropolitian-urbanized strata land domain index | N/A |
| RUCC2_sociod_EQI_22July2013 | non-metropolitian-urbanized strata sociodemographic domain index | N/A |
| RUCC2_built_EQI_22July2013 | non-metropolitian-urbanized strata built environment domain index | N/A |
| RUCC2_EQI_22July2013 | non-metropolitian-urbanized strata environmental quality index | N/A |
| RUCC3_air_EQI_22July2013 | less-urbanized strata air domain index | N/A |
| RUCC3_water_EQI_22July2013 | less-urbanized strata water domain index | N/A |
| RUCC3_land_EQI_22July2013 | less-urbanized strata land domain index | N/A |
| RUCC3_sociod_EQI_22July2013 | less-urbanized strata sociodemographic domain index | N/A |
| RUCC3_built_EQI_22July2013 | less-urbanized strata built environment domain index | N/A |
| RUCC3_EQI_22July2013 | less-urbanized strata environmental quality index | N/A |
| RUCC4_air_EQI_22July2013 | rural strata air domain index | N/A |
| RUCC4_water_EQI_22July2013 | rural strata water domain index | N/A |
| RUCC4_land_EQI_22July2013 | rural strata land domain index | N/A |
| RUCC4_sociod_EQI_22July2013 | rural strata sociodemographic domain index | N/A |
| RUCC4_built_EQI_22July2013 | rural strata built environment domain index | N/A |
| RUCC4_EQI_22July2013 | rural strata environmental quality index | N/A |
